# Trans-ancestry Genome-Wide Analyses in UK Biobank Yield Novel Risk Loci for Major Depression

**DOI:** 10.1101/2025.02.22.25322721

**Authors:** Madhurbain Singh, Chris Chatzinakos, Peter B. Barr, Amanda Elswick Gentry, Tim B. Bigdeli, Bradley T. Webb, Roseann E. Peterson

**Author notes:** Correspondence to <u>Madhurbain Singh</u>, Virginia Institute for Psychiatric and Behavioral Genetics, 800 E. Leigh St - Suite 100, Richmond, VA - 23298, USA.; <u>Roseann E. Peterson</u>, Institute for Genomics in Health, Department of Psychiatry and Behavioral Sciences, SUNY Downstate Health Sciences University, 450 Clarkson Ave, Brooklyn, NY 11203, USA.

## Abstract

Most genome-wide association studies (GWASs) of depression focus on broad, heterogeneous outcomes, limiting the discovery of genomic risk loci specific to major depressive disorder (MDD). Previous UK Biobank (UKB) studies had limited ability to pinpoint MDD-associated loci due to a smaller sample with strictly defined MDD outcomes and further exclusion of many participants based on ancestry or relatedness, significantly underutilizing this resource’s potential for elucidating the genetic architecture of MDD. Here, we present novel genomic insights into MDD by fully utilizing existing UKB data through (1) a trans-ancestry GWAS pipeline using two complementary approaches controlling for population structure and relatedness and (2) an increased sample with MDD symptom-level data across two mental health assessments. We identified strict MDD outcomes among 211,535 participants, representing a 38% increase in eligible participants from prior studies with only one assessment. Ancestrally inclusive analyses yielded 61 genomic risk loci across depression phenotypes, compared to 47 in the analyses restricted to participants genetically similar to European ancestry. Fourteen of these loci, including five novel, were associated with strict MDD phenotypes, whereas only one locus has been previously reported in UKB. MDD-associated genomic loci and predicted gene expression levels showed little overlap with broad depression, indicating higher specificity. Notably, polygenic scores based on these results were significantly associated with depression diagnoses across ancestry groups in the *All of Us* Research Program, highlighting the shared genetic architecture across populations. While the trans-ancestry analyses, which included non-European participants, increased the number of associated loci, the discovery of non-European ancestry-specific loci was limited, underscoring the need for larger, globally representative studies of MDD. Importantly, beyond these results, our GWAS pipeline will facilitate inclusive analyses of other traits and disorders, helping improve statistical power, representation, and generalizability in genomic studies.

## Introduction

Major depressive disorder (MDD) is a common, heterogeneous, and debilitating mental health problem and a leading cause of morbidity and mortality worldwide^1–3^. Twin and family studies indicate that additive genetic influences account for approximately 37% of the variance in MDD susceptibility^4^. Over the past decade, large-scale genome-wide association studies (GWASs) have provided valuable insights into the polygenic architecture of MDD and related depression outcomes^5–15^.

Genomic discovery for complex polygenic traits, like MDD, relies on ever-increasing sample sizes, typically achieved by meta-analyzing results across heterogeneously defined outcomes^9,10,13–15^. Structured clinical interviews are the gold standard for diagnosing MDD but have high financial and time costs^16^. Self-reported questionnaires with retrospective symptom-level information provide an alternative, relatively scalable approach to deriving strictly defined MDD phenotypes^16,17^. However, due to their wider availability, most data analyzed in the current large-scale studies^13–15^ come from broad, minimally defined depression phenotypes, such as electronic health records (EHR) and self-reported help-seeking or diagnosis. Prior research in UK Biobank^18^ (UKB) highlighted that GWAS of broad depression phenotypes primarily implicates genomic loci associated with general susceptibility to psychological distress rather than specific to MDD^17^.

In UKB, strict MDD phenotypes may be derived from self-reported retrospective symptoms assayed through the Composite International Diagnostic Interview Short Form (CIDI-SF)^19^ questionnaire. CIDI-SF was initially assessed in only around a third of the UKB participants (N∼153k in 2016–17). Previous GWASs were further restricted to unrelated participants either of self-reported White-British ethnicity^17^ or genetically similar to European ancestry^20^, thus excluding many potential participants due to genetic ancestry or relatedness. CIDI-SF data were reassessed in 2022-23 in a broader set of participants (N∼168k) but have yet to be analyzed in the GWAS of MDD. Extant GWASs in UKB have reported just one genomic risk locus for MDD^20^ and are yet to leverage this resource’s full potential for genomic discovery.

Furthermore, most data in current GWASs of depression come from participants genetically similar to European ancestry^9,13,21^. While strides have been made in increasing genetic diversity in studies of depression in recent years^10,11,14,15^, these studies meta-analyzed heterogeneous depression outcomes across available cohorts of different genetic ancestry groups. While we expect genetic architectures to be largely similar across populations, some genetic associations of complex traits may vary across ancestry groups due to differences in genetic background (e.g., genetic drift, allele frequencies, and linkage disequilibrium [LD]) and potential environmental moderators^21^. MDD and depression phenotypes show moderate genetic correlations across populations (e.g., 0.33–0.41 between European and East-Asian samples)^11,22^ and limited cross-population replicability of specific associations^11,14,22^. Thus, critical gaps remain in our understanding of MDD-specific genetic architecture across populations.

To address these limitations, we performed trans-ancestry GWAS of strictly defined MDD^17^ and broad depression^8^ outcomes in UKB (**Fig. 1A**). We assigned the UKB participants to ancestry groups based on genetic similarity to an expanded population reference panel. We combined the two CIDI-SF assessments to create a broader set of participants with strict MDD phenotypes. Building on the recommendations from the Psychiatric Genomics Consortium’s *Cross-Population Analyses Working Group*^21^, we employed two complementary approaches for conducting ancestrally inclusive GWAS: the *ancestry-stratified* meta-analysis and the *joint trans-ancestry* mixed-model GWAS. The results demonstrate the enhanced power achieved by this ancestrally inclusive GWAS pipeline, which revealed novel genomic loci associated with MDD and may facilitate additional trans-ancestry work in UKB and other datasets.

**Figure 1:**
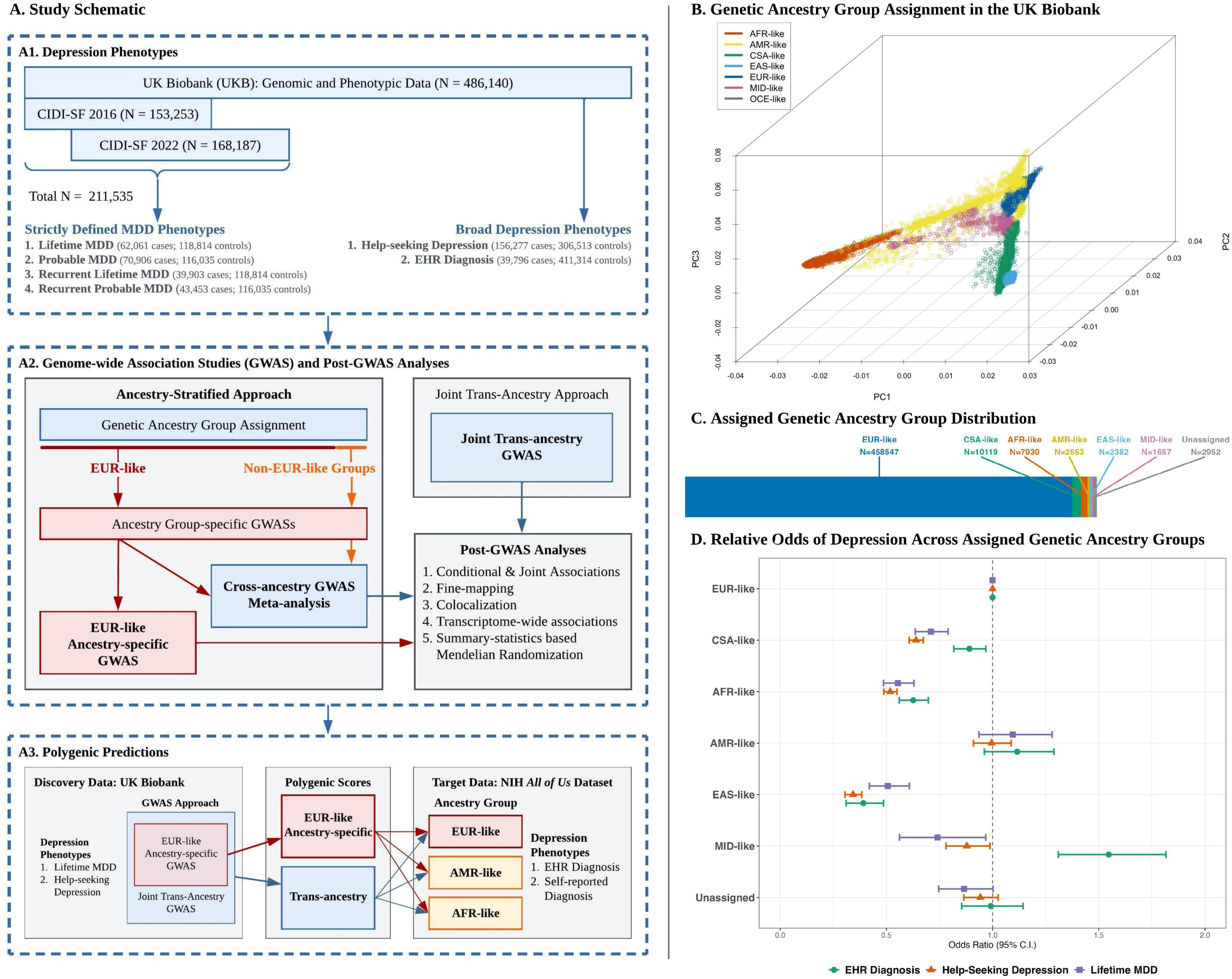
Study Overview, Genetic Ancestry Groups Assignment, and Differences in Depression Outcome Rates between Ancestry Groups. **A.** Study schematic presenting (**A1**) an overview of the depression outcomes, (**A2**) GWAS and post-GWAS analyses in UKB, and (**A3**) follow-up analyses in the All of Us Research Program. **B**. Genetic principal components analysis (PCA) plot showing the relative positions of UKB participants along the first three genetic PCs. Using PCA, the participants were assigned to “genetic ancestry groups” based on their genetic similarity to major global populations in the 1000 Genomes Project Phase-3 and Human Genome Diversity Project: European/EUR-like, Central and South Asian/CSA-like, African/AFR-like, Admixed American/AMR-like, East Asian/EAS-like, and Middle Eastern MID-like. **C**. Sample size distribution of the assigned genetic ancestry groups. **D**. Relative odds of different depression outcomes in the non-EUR-like groups relative to the EUR-like group, controlling for age, sex, and UKB assessment center.

## Methods

### Depression Phenotypes in UK Biobank

UKB is an ongoing population-based prospective study of ∼500,000 participants from the UK, aged 40-69 at recruitment (2006–2010), with rich phenotypic, genetic, and linked EHR data^18^. We operationalized strict MDD phenotypes in UKB based on self-reported retrospective symptoms aggregated across the two CIDI-SF^19^ assessments in partially overlapping subsets of participants (**Supplementary Methods**, **Supplementary Table 1**). Building on prior research^17^, *LifetimeMDD* cases were defined as those meeting the DSM-5 (Diagnostic and Statistical Manual of Mental Disorders, Fifth Edition)^23^ MDD criteria of symptoms, episode length, and accompanying distress or functional impairment. We additionally derived *ProbableMDD* cases requiring at least four self-reported MDD symptoms (compared to five symptoms in DSM-5), reflecting the CIDI-SF criterion for epidemiological screening of MDD^19^. Further, recurrent cases of both phenotypes (*RecLifetimeMDD*, *RecProbableMDD*) were identified based on the reported lifetime number of such episodes. For comparison with prior studies^8,17^ using broad depression outcomes assessed in nearly all participants, we defined Help-seeking Depression (*HelpSeekingDep*) based on participants’ self-reports of seeking medical help for psychological distress symptoms. *EHR Diagnosis* (*EHRdep*) was derived by aggregating depression-related diagnostic entries (primary or secondary) across primary care, hospital inpatient, and death register records.

### Genetic Similarity-Based Ancestry Group Assignment

For ancestry-stratified analyses, we used the *POP-MaD* (population grouping by Mahalanobis distance) pipeline^24^ to assign the UKB participants to ancestry groups^25^ based on genetic similarity to seven global population groups in the 1000 Genomes Project Phase-3 and the Human Genome Diversity Project^26^ (African/AFR, Admixed American/AMR, Central and South Asian/CSA, East Asian/EAS, European/EUR, Middle-Eastern/MID, and Oceania/OCE; **Supplementary Methods**). The derived groups were assigned labels AFR-like, AMR-like, CSA-like, EAS-like, EUR-like, MID-like, and OCE-like, reflecting the label of the reference population with which the group clustered. Potential “outliers” (Mahalanobis distance more than three standard deviations from the group median) were excluded from the ancestry-stratified analyses to further reduce within-group genetic heterogeneity.

### Association Analyses

For the ancestry-stratified meta-analysis^21^ (**Fig. 1**, **Panel A2**), we first performed genetic ancestry group-specific GWASs (including the EUR-like ancestry-specific GWAS typically conducted in UKB), followed by a cross-ancestry fixed-effect meta-analysis^27^ (**Supplementary Methods**). In the joint trans-ancestry approach, the GWAS is conducted across all participants using linear mixed models (LMMs) to account for genetic similarities between participants. In both approaches, we performed GWAS using REGENIE^28^, which implements a whole-genome regression model closely related to the LMM and helps to control for population structure and genetic relatedness between samples. Thus, we also included related samples in both the ancestry-stratified and the joint trans-ancestry GWAS. For each set of results, independent genome-wide significant (GWS; *p* < 5 × 10^−8^) single nucleotide polymorphisms (SNPs) were identified using conditional-and-joint GCTA-COJO^29^ analyses, and the overlap of GWS loci (COJO-index SNP ±250kb) was then examined between phenotypes and analytical approaches (**Supplementary Methods**).

### SNP Heritability and Genetic Correlations

We used linkage disequilibrium score regression (LDSC)^30^ to estimate the SNP-based heritability (phenotypic variance explained by genome-wide SNPs) of each depression phenotype and the genetic correlations (*r*_*G*_) between phenotypes (**Supplementary Methods**). As LDSC requires relatively homogeneous ancestry samples, these analyses were limited to the EUR-like ancestry-specific GWAS, as the other groups were not large enough for ancestry-specific post-GWAS analyses.

### Fine-mapping and Colocalization

At each genomic locus associated with any of the six depression phenotypes, we performed statistical fine-mapping (using *SuSieR*^31^) to identify credible sets of SNPs likely driving the locus association (**Supplementary Methods**). We then performed colocalization analyses (using *coloc*^32^) to examine whether the different phenotypes share putative causal SNPs at each locus, even if the locus does not reach the GWS threshold for a phenotype in GWAS.

### Post-GWAS Gene-Level Analyses in the Brain

We performed transcriptome-wide association study (TWAS)^33^ analyses using *JEPEGMIX2-P*^34^ to predict brain region-specific gene expression levels associated with depression phenotypes, leveraging external data on expression quantitative loci (eQTL) in 13 brain regions from GTEx^35^ (**Supplementary Methods**). Further, we performed summary-statistics-based Mendelian Randomization (SMR^36^) analyses with the GWAS associations and the eQTL data to examine whether gene expression levels in these brain regions putatively mediate the SNP-depression association (**Supplementary Methods**). In both analyses, a within-tissue false discovery rate <0.05 was considered statistically significant after multiple-testing correction. As well-powered eQTL data were only available for EUR-like ancestry groups, these analyses were limited to the EUR-like ancestry-specific GWAS^37^.

### Polygenic Score Testing in the *All of Us* Research Program

To test the individual-level replicability and cross-ancestry transferability of the genetic associations discovered in UKB, we tested polygenic scores (PGSs) in the *All of Us* Research Program (AoU)^38^. We derived PGSs based on EUR-like ancestry-specific and joint trans-ancestry GWAS summary statistics of *LifetimeMDD* and *HelpSeekingDep*, using PRS-CS^39^ (**Supplementary Methods**). PGS associations with EHR-based and self-reported depression diagnoses in the EUR-like (N=112,005), AFR-like (N=41,149), and AMR-like (N=32,958) unrelated subsets of AoU participants were estimated using logistic regression, controlling for age, gender, and the first 10 genetic principal components (PCs).

## Results

### Extended Sample with Strictly Defined MDD Phenotypes

We identified 62,061 cases and 118,814 controls of *LifetimeMDD* (**Fig 1A**) out of 211,535 participants with at least one assessment of self-reported MDD symptoms across the two CIDI-SF surveys (**Supplementary Table 2**). This aggregated study sample yielded a 38% increase in eligible participants for GWAS of MDD, compared to N∼153k participants evaluated in the first assessment, of which previous studies examined ∼67k (self-identified “White-British”)^17^ to ∼93k (EUR-like genetic ancestry)^20^ unrelated participants.

### Expanded Genetic Ancestry Group Assignments

Updated genetic ancestry group assignment using *POP-MaD* yielded subsets of 458,547 EUR-like, 10,119 CSA-like, 7,930 AFR-like, 2,553 AMR-like, 2,382 EAS-like, and 1,657 MID-like participants, representing >5% non-EUR-like participants, while 2,952 participants were classified as ancestry-group “outliers” (**Figures 1B-C**, **Supplementary Tables 3**, **Supplementary Figure 1**). Compared to previous assignments based on a random-forest algorithm used in the “Pan-UKBB” initiative^40^, *POP-MaD* increased the available sample size by 36,621 (8.2%) for ancestry-stratified meta-analysis, including 33,069 (7.9%) for EUR-like ancestry-specific analyses (**Supplementary Tables 4**). The genetic ancestry groups showed significant mean differences in case prevalences of both strict MDD and broad depression outcomes (**Fig. 1D**, **Supplementary Table 5**), with CSA-like, AFR-like, EAS-like, and MID-like participants being less likely to meet the case criteria than EUR-like participants.

Importantly, it should be noted that genetic similarity-based subsets are created to reduce genetic heterogeneity for statistical analyses (e.g., to minimize population stratification) and should not be considered synonymous with social constructs of racial and ethnic identities^25^. In the joint trans-ancestry analyses, all participants were included without considering such groupings while statistically accounting for their genetic similarity.

### Inclusive GWAS Approaches Maximize the Power for Genomic Discovery and Uncover Novel Loci for MDD

We identified genome-wide significant (GWS; *p* < 5 × 10^−8^) genetic variants in three sets of results: EUR-like ancestry-specific GWAS, cross-ancestry meta-analysis, and joint trans-ancestry GWAS. Ancestry-specific GWASs in CSA-like, AFR-like, AMR-like, EAS-like, and MID-like subsets did not yield any GWS loci, likely because the sample sizes were <1-2% of the EUR-like group. In total, these analyses identified 61 independent genomic loci across depression phenotypes (**Fig. 2**, **Supplementary Tables 6-12**, **Supplementary Figure 2**). Including non-EUR-like participants (via cross-ancestry (*stratified*) and trans-ancestry (*joint*) GWAS approaches) resulted in 14 additional GWS loci despite a modest sample size increase of ∼6%. Conversely, three loci in the EUR-like ancestry group-specific analyses were no longer significant in the cross-ancestry GWASs, though with largely overlapping confidence intervals across approaches. Fourteen loci (including five novel loci; **Table 1**, **Supplementary Figures 3-4**) were associated with one or more strict MDD phenotypes, compared to only one locus previously reported for *LifetimeMDD* in the unrelated EUR-like subset^20^. Consistent with prior findings, the highest number of GWS loci (48) were identified in the analyses of *HelpSeekingDep*, likely reflecting its largest sample size.

**Figure 2:**
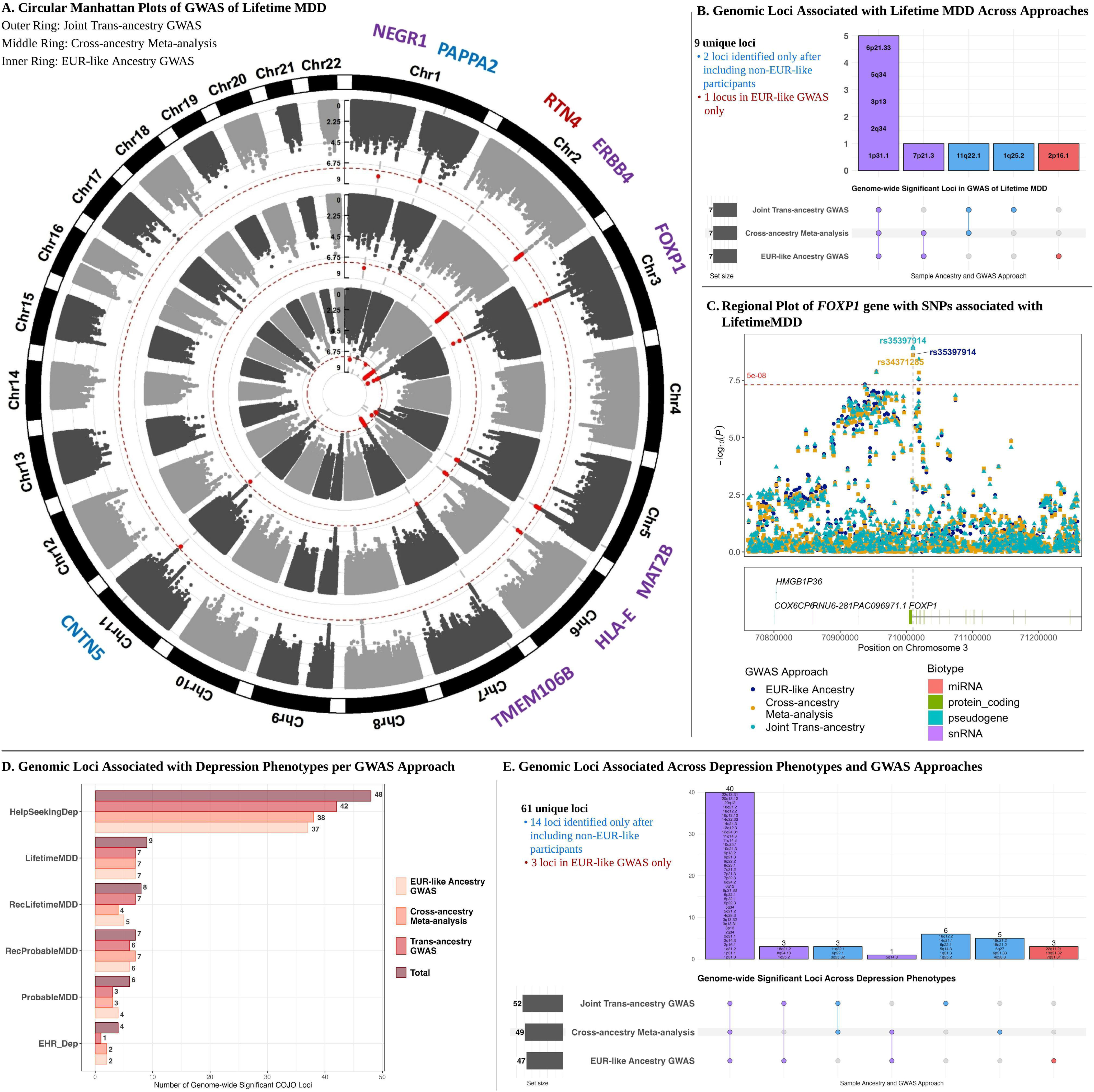
Genomic Loci Associated with Lifetime MDD and Other Depression Phenotypes across GWAS Approaches. **A.** Circular Manhattan plots of the GWAS of Lifetime MDD, with outer to inner rings showing the joint trans-ancestry GWAS, the cross-ancestry GWAS meta-analysis, and the EUR-like ancestry-specific GWAS, respectively. The X-axis shows the positions of the SNPs along the chromosomes, and the Y-axis shows the respective -log10(p-value). The SNPs meeting the genome-wide significance (GWS) threshold (*p* < 5 × 10^−8^) are shown in red. The labels along the circumference are the nearest mapping genes of the GWS loci. **B.** Upset plot showing the overlap of GWS loci across the three GWAS results of Lifetime MDD. **C.** Regional plot centered at the index SNPs in *FOXP1* (at 3p13), one of the novel risk loci for Lifetime MDD. The X-axis shows the SNP positions on chromosome 3 (aligned to GRCh37), and the Y-axis shows the -log10(p-value). The estimates from the three GWASs are shown in different colors and overlaid on each other. **D.** Number of independent genomic loci associated at the GWS level with each depression phenotype across different GWAS approaches. **E.** Upset plot showing the overlap of GWS loci between the three main sets of GWAS results, pooled across depression phenotypes. The matrix rows represent GWAS approaches, with the horizontal bars showing the number of GWS loci per approach. The columns represent the overlap between the indicated approaches, with the corresponding vertical bar depicting the number of overlapping loci.

**Table 1.**
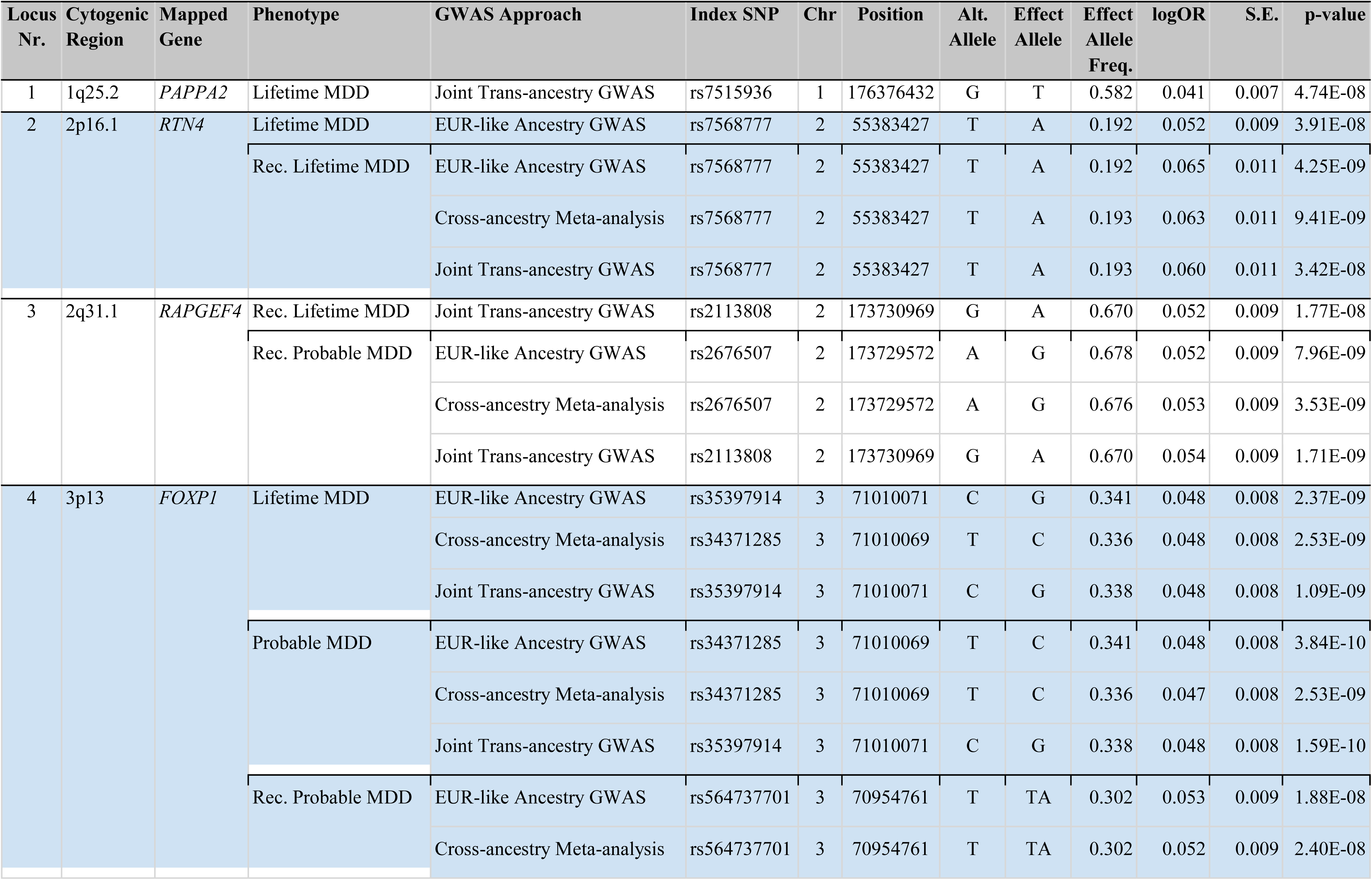
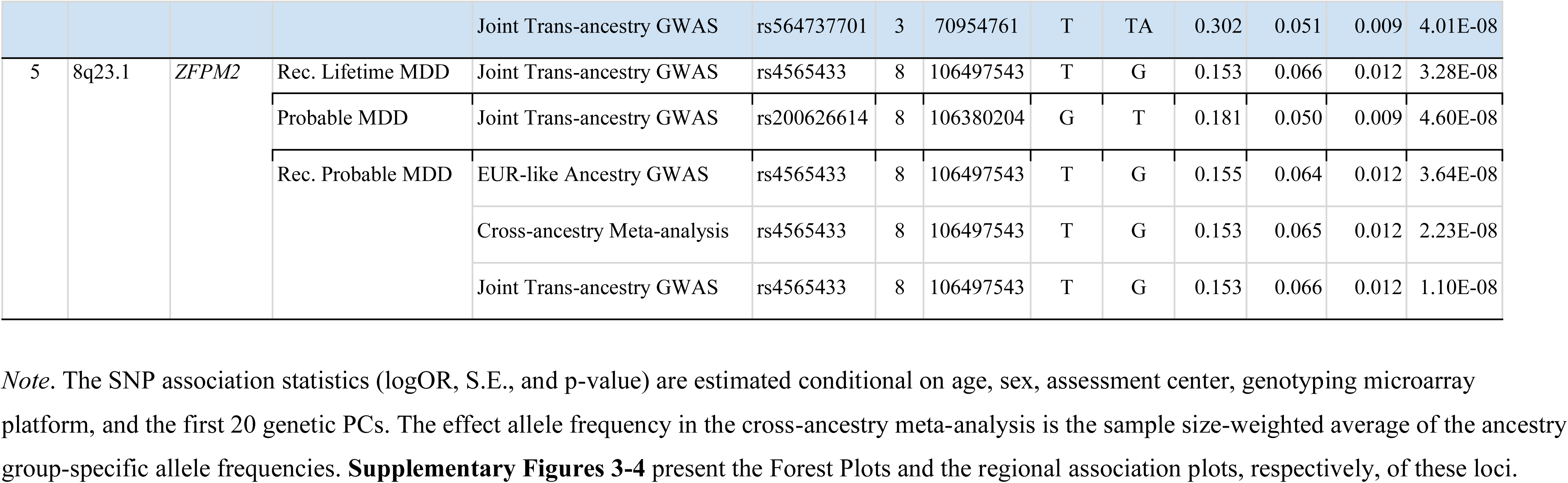
Novel Risk Loci Genome-wide Significantly Associated with Strictly Defined MDD Phenotypes.

| Locus Nr. | Cytogenic Region | Mapped Gene | Phenotype | GWAS Approach | Index SNP | Chr | Position | Alt. Allele | Effect Allele | Effect Allele Freq. | logOR | S.E. | p-value |
| --- | --- | --- | --- | --- | --- | --- | --- | --- | --- | --- | --- | --- | --- |
| 1 | 1q25.2 | PAPPA2 | Lifetime MDD | Joint Trans-ancestry GWAS | rs7515936 | 1 | 176376432 | G | T | 0.582 | 0.041 | 0.007 | 4.74E-08 |
| 2 | 2p16.1 | RTN4 | Lifetime MDD | EUR-like Ancestry GWAS | rs7568777 | 2 | 55383427 | T | A | 0.192 | 0.052 | 0.009 | 3.91E-08 |
|  |  |  | Rec. Lifetime MDD | EUR-like Ancestry GWAS | rs7568777 | 2 | 55383427 | T | A | 0.192 | 0.065 | 0.011 | 4.25E-09 |
|  |  |  |  | Cross-ancestry Meta-analysis | rs7568777 | 2 | 55383427 | T | A | 0.193 | 0.063 | 0.011 | 9.41E-09 |
|  |  |  |  | Joint Trans-ancestry GWAS | rs7568777 | 2 | 55383427 | T | A | 0.193 | 0.060 | 0.011 | 3.42E-08 |
| 3 | 2q31.1 | RAPGEF4 | Rec. Lifetime MDD | Joint Trans-ancestry GWAS | rs2113808 | 2 | 173730969 | G | A | 0.670 | 0.052 | 0.009 | 1.77E-08 |
|  |  |  | Rec. Probable MDD | EUR-like Ancestry GWAS | rs2676507 | 2 | 173729572 | A | G | 0.678 | 0.052 | 0.009 | 7.96E-09 |
|  |  |  |  | Cross-ancestry Meta-analysis | rs2676507 | 2 | 173729572 | A | G | 0.676 | 0.053 | 0.009 | 3.53E-09 |
|  |  |  |  | Joint Trans-ancestry GWAS | rs2113808 | 2 | 173730969 | G | A | 0.670 | 0.054 | 0.009 | 1.71E-09 |
| 4 | 3p13 | FOXP1 | Lifetime MDD | EUR-like Ancestry GWAS | rs35397914 | 3 | 71010071 | C | G | 0.341 | 0.048 | 0.008 | 2.37E-09 |
|  |  |  |  | Cross-ancestry Meta-analysis | rs34371285 | 3 | 71010069 | T | C | 0.336 | 0.048 | 0.008 | 2.53E-09 |
|  |  |  |  | Joint Trans-ancestry GWAS | rs35397914 | 3 | 71010071 | C | G | 0.338 | 0.048 | 0.008 | 1.09E-09 |
|  |  |  | Probable MDD | EUR-like Ancestry GWAS | rs34371285 | 3 | 71010069 | T | C | 0.341 | 0.048 | 0.008 | 3.84E-10 |
|  |  |  |  | Cross-ancestry Meta-analysis | rs34371285 | 3 | 71010069 | T | C | 0.336 | 0.047 | 0.008 | 2.53E-09 |
|  |  |  |  | Joint Trans-ancestry GWAS | rs35397914 | 3 | 71010071 | C | G | 0.338 | 0.048 | 0.008 | 1.59E-10 |
|  |  |  | Rec. Probable MDD | EUR-like Ancestry GWAS | rs564737701 | 3 | 70954761 | T | TA | 0.302 | 0.053 | 0.009 | 1.88E-08 |
|  |  |  |  | Cross-ancestry Meta-analysis | rs564737701 | 3 | 70954761 | T | TA | 0.302 | 0.052 | 0.009 | 2.40E-08 |
|  |  |  |  | Joint Trans-ancestry GWAS | rs564737701 | 3 | 70954761 | T | TA | 0.302 | 0.051 | 0.009 | 4.01E-08 |
| 5 | 8q23.1 | ZFPM2 | Rec. Lifetime MDD | Joint Trans-ancestry GWAS | rs4565433 | 8 | 106497543 | T | G | 0.153 | 0.066 | 0.012 | 3.28E-08 |
|  |  |  | Probable MDD | Joint Trans-ancestry GWAS | rs200626614 | 8 | 106380204 | G | T | 0.181 | 0.050 | 0.009 | 4.60E-08 |
|  |  |  | Rec. Probable MDD | EUR-like Ancestry GWAS | rs4565433 | 8 | 106497543 | T | G | 0.155 | 0.064 | 0.012 | 3.64E-08 |
|  |  |  |  | Cross-ancestry Meta-analysis | rs4565433 | 8 | 106497543 | T | G | 0.153 | 0.065 | 0.012 | 2.23E-08 |
|  |  |  |  | Joint Trans-ancestry GWAS | rs4565433 | 8 | 106497543 | T | G | 0.153 | 0.066 | 0.012 | 1.10E-08 |

*Note.* The SNP association statistics (logOR, S.E., and p-value) are estimated conditional on age, sex, assessment center, genotyping microarray platform, and the first 20 genetic PCs. The effect allele frequency in the cross-ancestry meta-analysis is the sample size-weighted average of the ancestry group-specific allele frequencies. **Supplementary Figures 3-4** present the Forest Plots and the regional association plots, respectively, of these loci.

To examine potential confounding due to relatedness or population structure, we compared the LDSC intercepts of GWASs performed as sensitivity analyses in the EUR-like subset by excluding related participants or ancestry-group “outliers” defined with varying stringency (**Supplementary Methods**). Notably, these analyses showed negligible non-systematic differences in their LDSC intercept, supporting the robustness of GWAS to these sources of confounding (**Supplementary Table 13**, **Supplementary Figure 5**).

For *LifetimeMDD*, nine significant loci, including three novel (**Table 1**), were identified. Two loci were discovered only upon including non-EUR-like participants (**Fig. 2B**), comprising one novel (rs7515936 upstream of *PAPPA2*) and one previously reported (rs4388849 in *CNTN5*)^14^ locus. One novel locus (rs7568777 upstream of *RTN4*) showed a significant association with *LifetimeMDD* in the EUR-like subset only but was associated with *RecLifetimeMDD* in all three GWAS approaches. Another novel locus in *FOXP1* (**Fig. 2C**) had intronic SNPs associated with *LifetimeMDD*, *RecLifetimeMDD*, and *RecProbableMDD* in all three approaches. The remaining two novel MDD loci were tagged by intronic index SNPs in *RAPGEF4* and *ZFPM2* and associated with recurrent MDD phenotypes.

Ancestrally inclusive GWAS approaches also improved fine-mapping resolution at the associated loci (**Supplementary Tables 14-19**). Notably, the ancestry-stratified meta-analysis yielded the smallest median credible set size (fewest fine-mapped SNPs per credible set) and identified the most sets containing higher confidence fine-mapped SNPs with a posterior inclusion probability >0.50 (**Supplementary Figure 6**).

### Strict MDD Phenotypes are Associated with Genomic Loci Not Shared with Broad Depression

Consistent with prior literature^17^, strict MDD phenotypes had higher SNP-based heritability than broad depression ones, ranging from 0.074 (S.E. 0.005) for *EHRdep* to 0.133 (S.E. 0.008) for *RecLifetimeMDD* in the EUR-like subset (Fig. 3A, **Supplementary Tables 20-21**). The four MDD phenotypes showed near-perfect *r*_*G*_ with each other (range: 0.99–1.00; in the EUR-like subset) but slightly lower *r*_*G*_ with *HelpSeekingDep* (0.87–0.89) and lower still with *EHRdep* (0.81–0.82), indicating that MDD shares approximately 79% (*r*^2^ × 100) genetic variance with *HelpSeekingDep* and 67% with *EHRdep* (Fig. 3B). Of the 14 genomic risk loci identified across strict MDD phenotypes, only four loci were also GWS with *HelpSeekingDep* or *EHRdep* (despite their much larger sample sizes), suggesting higher specificity for MDD (Fig. 3C). Importantly, none of the five novel MDD loci were GWS in the analyses of broad depression phenotypes. In colocalization analyses applied across all depression-associated loci in each GWAS approach, the MDD phenotypes shared putative causal SNPs at 35–44% of the loci (Fig. 3D, **Supplementary Tables 22-25**). In contrast, the MDD phenotypes shared putative causal SNPs with *HelpSeekingDep* at 20–22% of loci and *EHRdep* at only 7–13%.

**Figure 3:**
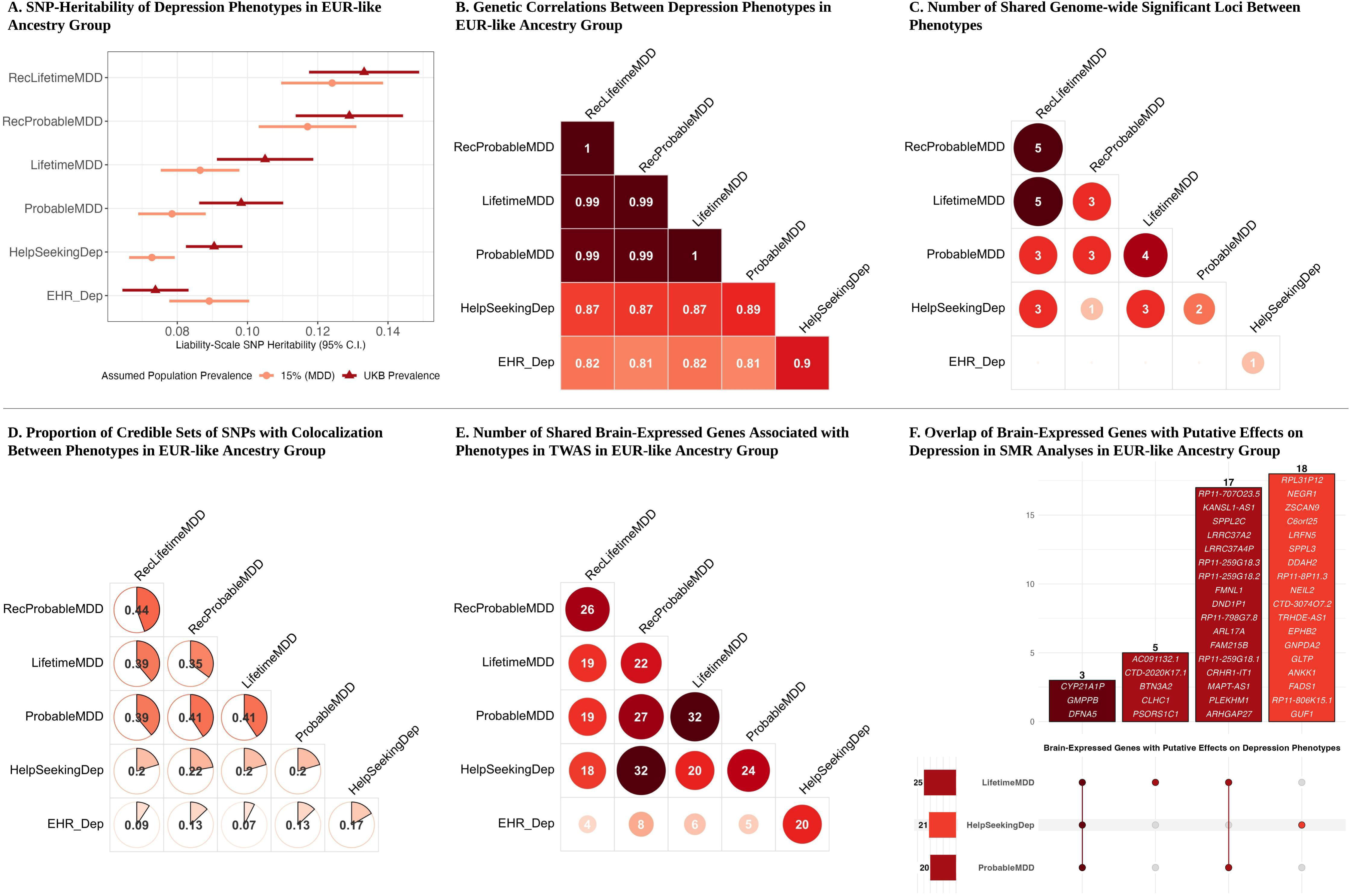
Overlap between Depression Phenotypes at Genome-wide and Locus-level Resolution. **A.** Genome-wide SNP-based heritabilities in the EUR-like subset estimated using LDSC analyses. The plot shows the point estimate and the 95% confidence interval. Two sets of liability-scale heritabilities are shown, with different assumed population prevalence rates of the depression outcomes: 15% (prevalence of MDD) and the phenotype-specific overall prevalence observed in UKB. **B.** Genome-wide SNP-based genetic correlations in the EUR-like subset estimated using LDSC analyses and assuming UKB prevalence to be the population prevalence rate. **C.** Number of genome-wide significant risk loci overlapping between phenotypes. The number of loci per phenotype is aggregated across the three GWAS results and then examined for overlap with the other phenotypes. **D.** Proportion of genomic risk loci showing significant (pair-wise) colocalization between phenotypes based on the EUR-like ancestry-specific GWAS. Similar overlaps were seen with the other GWAS results (**Supplementary Table 25**). **E.** Number of overlapping unique genes (in 13 brain regions) significantly associated with different phenotypes in the EUR-like ancestry-specific TWAS. **F.** Upset plot showing the overlap of unique genes (across 13 brain regions) with significant putative causal effects on depression phenotypes in the EUR-like ancestry-specific SMR analyses. The matrix rows represent depression phenotypes, with the corresponding horizontal bar showing the number of causally linked genes. The columns represent the overlap between the indicated phenotypes, with the corresponding vertical bar showing the number and names of the overlapping genes.

### HPA Axis Genes are Implicated in Lifetime MDD but not Broad Depression

TWAS analyses revealed 299 unique genes with brain region-specific expression levels associated with the six depression phenotypes, of which 78 were associated with one or more strict MDD phenotypes (**Supplementary Tables 26-31**). Consistent with the GWAS results, *HelpSeekingDep* was associated with considerably more genes (244) than the other phenotypes: 44 with *LifetimeMDD*, 47 with *ProbableMDD*, 33 with *RecLifetimeMDD*, 46 with *RecProbableMDD*, and 34 with *EHRdep*. The MDD phenotypes shared around half of the associated genes with each other and with *HelpSeekingDep* (Fig. 3E), indicating unique gene-level associations discovered with strict MDD phenotypes despite their smaller sample size. To illustrate, only 20 genes were shared between *LifetimeMDD* and *HelpSeekingDep* (Fig. 4A).

**Figure 4:**
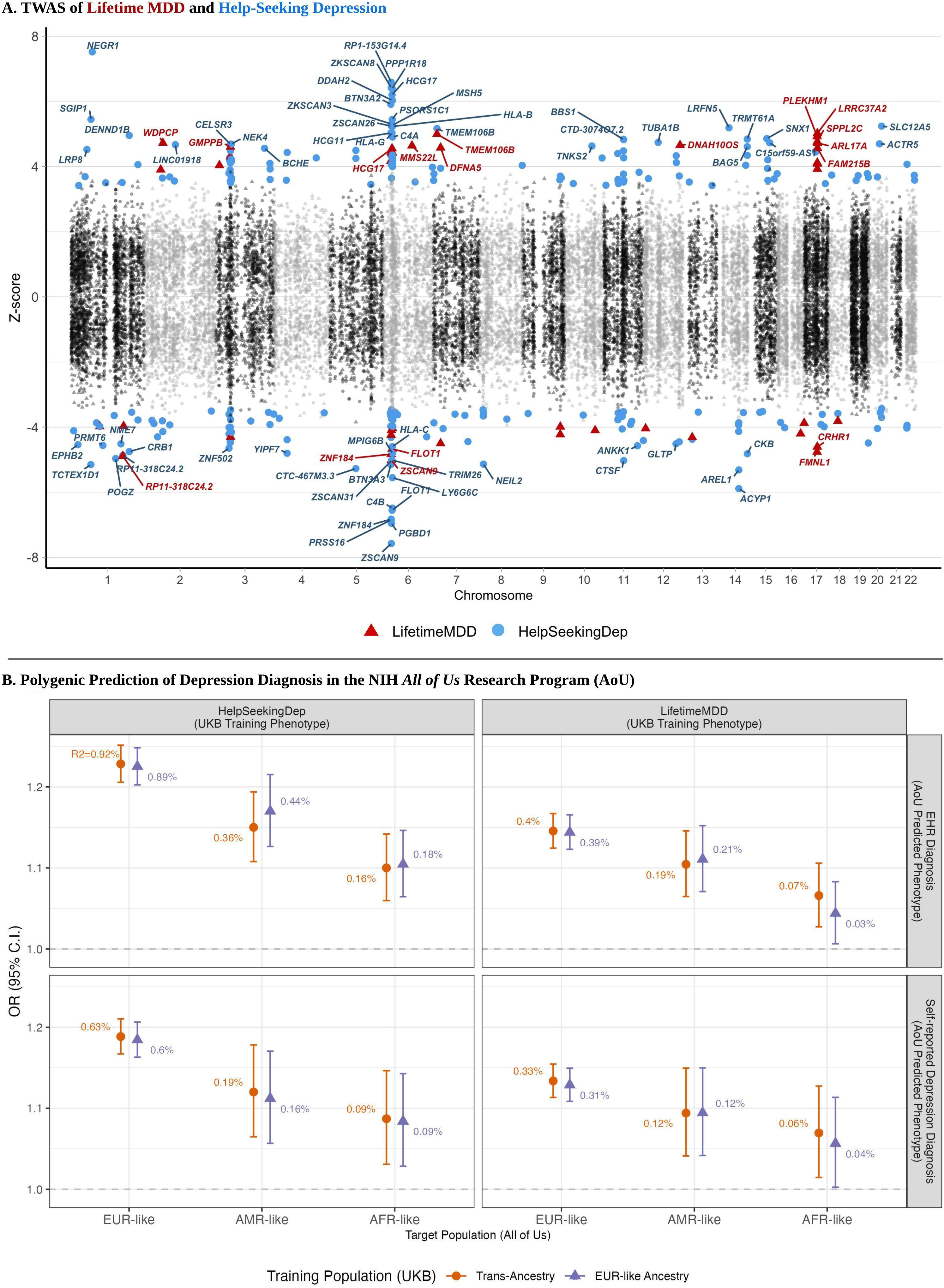
Follow-up Analyses Comparing Lifetime MDD and Help-Seeking Depression: TWAS in the Brain and Polygenic Prediction. **A.** Miami plot showing genes predicted to be associated with Lifetime MDD and Help-Seeking Depression in TWAS analyses based on the EUR-like ancestry-specific GWAS. TWAS analyses were performed across 13 brain regions. The Y-axis indicates each gene’s Z-score in the brain region with the most significant association with each phenotype. Significant associations with Lifetime MDD are shown in red, while those with Help-Seeking Depression are shown in blue. **B.** Associations of UKB-based PGSs of Lifetime MDD and Help-Seeking Depression with depression diagnoses in the EUR-like, AMR-like, and AFR-like participants from the All of Us Research Program (AoU). The vertical panels indicate the UKB depression phenotype used to compute the PGSs, and the horizontal panels indicate the depression diagnosis being predicted in AoU (EHR-based and self-reported). The error bars indicate the 95% confidence interval of the odds ratio, and the text label indicates the incremental pseudo-R^2^ (in %). All PGS associations are conditional on age, gender, and the first 10 genetic PCs and are statistically significant at FDR <0.05. The PGSs shown here were based on the EUR-like ancestry-specific and the joint trans-ancestry GWASs. **Supplementary Figure 8** compares these results with PGSs computed through cross-ancestry meta-analysis in PRS-CSx.

Here, two notable clusters of genes in high-LD chromosomal regions emerged: the genes in the major histocompatibility complex region associated with both *LifetimeMDD* and *HelpSeekingDep*, and the genes at 17q21.31 associated almost exclusively with *LifetimeMDD*. The latter region includes genes involved in the HPA (hypothalamus-pituitary-adrenal) axis and stress response, including the *CRHR1* (corticotropin-releasing hormone receptor 1) gene, not associated with *HelpSeekingDep*. The 17q21.31 locus contains a common inversion polymorphism in EUR-like populations^41^, and the mapped gene cluster has previously been implicated in post-traumatic stress disorder^42^, autism spectrum disorder^43^, and neurodegenerative disorders^41^.

Further SMR analyses showed significant putative causal influences of brain region-specific gene expression levels on *LifetimeMDD*, *ProbableMDD*, and *HelpSeekingDep* (Fig. 3F, **Supplementary Tables 32-34**). The 20 genes putatively influencing *ProbableMDD* were all a subset of the 25 genes influencing *LifetimeMDD*. However, of the 21 genes with potential effects on *HelpSeekingDep*, only three were shared with *LifetimeMDD*. Thus, these analyses highlighted genes likely specific to MDD (e.g., *CRHR1-IT1* and *LRRC37A4P* [17q21.31]) versus those shared with *HelpSeekingDep* (e.g., *GMPPB* and *DFNA5*).

### Polygenic Scores from UKB Significantly Associated with Depression Diagnoses Across Ancestry Groups in AoU

Using EUR-like ancestry-specific GWAS summary statistics from UKB, both *LifetimeMDD* and *HelpSeekingDep* PGSs were significantly associated with EHR-derived and self-reported depression diagnoses in the EUR-like subset of AoU participants (Fig. 4B, **Supplementary Table 35**). The *HelpSeekingDep* PGS had a stronger association with depression diagnoses (O.R. = 1.23; 95% confidence interval (C.I.) = [1.20, 1.25]; incremental R^2^ = 0.89% for EHR Diagnosis) than *LifetimeMDD* PGS (O.R. = 1.14; C.I. = [1.12, 1.17]; incremental R^2^ = 0.39%), which prior research^17^ has demonstrated to be due to the former’s larger GWAS sample size. The association of PGSs based on trans-ancestry GWAS results showed trivial, non-significant differences from EUR-like ancestry-based PGSs, likely because EUR-like participants comprised ∼95% of the UKB trans-ancestry sample. Notably, both EUR-like ancestry-based and trans-ancestry PGSs were also significantly associated with depression diagnoses in the AMR-like and AFR-like participants, albeit with reduced effect sizes. For instance, for the association of EUR-like ancestry-based *HelpSeekingDep* PGS with EHR depression diagnosis: O.R. = 1.17; C.I. = [1.13, 1.22]; incremental R^2^ = 0.44% in AMR-like participants, and O.R. = 1.10; C.I. = [1.06, 1.15]; incremental R^2^ = 0.18% in AFR-like participants.

## Discussion

This study presents novel MDD-specific genomic insights in UKB through two key innovations: a considerably larger set of participants with strictly defined MDD phenotypes from two repeated assessments of symptom-level data and an inclusive, trans-ancestry GWAS pipeline accommodating related participants across the genetic ancestry continuum, which maximized the statistical power for genomic discovery. While prior UKB analyses showed that broad depression outcomes are primarily associated with genomic loci not specific to MDD^17^, previous studies used smaller samples and could identify just one MDD-associated locus. Here, we identified 14 independent genomic risk loci, including five novel, associated with MDD phenotypes. The MDD-associated genomic loci and TWAS-predicted gene expression levels in the brain showed little overlap with broad depression, indicating higher specificity. Despite 95% of UKB participants being genetically similar to EUR ancestry, the trans-ancestry analyses uncovered 14 additional genomic loci across depression phenotypes, highlighting the value of ancestrally inclusive GWAS. This pipeline implements the ethical imperative of adopting more inclusive GWAS approaches, ensuring that, as far as possible, the analyses include all participants who contributed DNA and survey data.

Two of the five novel genomic risk loci for MDD are intergenic regions upstream of *PAPPA2* and *RTN4*. *PAPPA2* regulates the bioavailability of insulin-like growth factor^44^, a neurotrophic hormone crucial for central nervous system development, maturation, and neuroplasticity^45^. *RTN4* inhibits neurite outgrowth and axon regeneration and plasticity^46^. Notably, high *RTN4* expression levels are postulated as a state-level blood biomarker of psychological stress^47^ and hallucinations^48^ and predict future hospitalizations attributed to these symptoms^47,48^. The other three loci are intronic regions in *RAPGEF4*, *FOXP1*, and *ZFPM2*. *RAPGEF4* is expressed primarily in the brain, neuroendocrine, and endocrine tissues^49^ and is involved in neuronal differentiation, neurite outgrowth, neurotransmitter release, memory, and learning^49^, as well as corticotropin-releasing hormone-mediated dendritic spine loss in acute stress^50^. Moreover, *RAPGEF4* knock-out mice show depression- and anxiety-like behaviors responsive to fluoxetine (selective serotonin reuptake inhibitor)^51^. *FOXP1* encodes a transcription factor critical to the early development of several organ systems, including the brain^52^. Deletions or mutations in this gene cause *FOXP1 Syndrome*, characterized by, among other features, language deficits, autism spectrum disorder, intellectual disability, and congenital brain anomalies^52^. *ZFPM2* is a zinc-finger transcription factor associated with GABAergic neuron differentiation in the midbrain and hindbrain tegmental nuclei that control mood, memory, motivated behavior, and movement^53^.

These novel loci have previously been reported to be linked with other MDD-associated traits. For instance, *PAPPA2*, *RTN4*, *FOXP1*, and *ZFPM2* are associated with body weight^54^, body mass index^55^, or body fat percentage^56^. *RTN4*, *RAPGEF4*, *FOXP1*, and *ZFPM2* are associated with educational attainment^57^ (in the opposite direction of effect). *RTN4*, *RAPGEF4*, and *FOXP1* are associated with substance use^58^ or other risk-taking/externalizing behaviors^59^. Notably, SNPs in *FOXP1* have also been implicated in GWASs of retrospectively self-reported childhood maltreatment^60^, insomnia^61^, and neuroimaging-based total brain volume^62^ and sulcal depth^63^. These pleiotropic associations may arise from horizontal pleiotropy due to shared endophenotypes and/or vertical pleiotropy due to causal influences between MDD and other traits.

PGSs based on UKB GWAS results showed significant within- and cross-ancestry associations with depression diagnoses in AoU, highlighting the shared genetic architecture of MDD across populations. However, consistent with an increasing genetic distance from EUR ancestry, the effect sizes reduced progressively in AMR-like and AFR-like participants, underscoring the need for globally representative genetic studies to prevent exacerbating health disparities. The diminished associations may also reflect differences in healthcare access and, thus, depression diagnosis between populations, further reducing the transferability of UKB GWAS results to non-EUR-like participants.

When interpreting these findings, it should be noted that the MDD phenotypes were not obtained from gold-standard clinical diagnostic interviews. Although phenotypes based on structured clinical interviews and self-reported retrospective symptoms from CIDI-SF likely lie on the same genetic liability continuum^17^, the respondent’s current mood may prime the recall of past episodes in the latter. Secondly, prior research has shown that participation in the online mental health questionnaire in UKB is genetically correlated with several mental health traits^64^, though follow-up analyses found no evidence of its influence on MDD case-control status^17^. UKB is also enriched for healthy volunteers with overall better health and higher levels of education than the general UK population^65^, which could introduce bias in GWAS of behavioral traits^66^.

In this study, we performed both ancestry-stratified meta-analysis and joint trans-ancestry GWAS, following best practices^21^. The ancestry group-specific analyses in the former exclude the participants identified as potential ancestry group outliers but can facilitate post-GWAS analyses requiring ancestrally matched external reference data, such as LDSC analyses^30^ for SNP-based heritability and genetic correlations, gene-level TWAS^33^ and SMR^36^ analyses, and PGS analyses^39^. In comparison, the joint trans-ancestry approach is the most inclusive as it includes all participants across the genetic ancestry continuum. Given the predominance of EUR-like participants in UKB, the trans-ancestry genetic analyses in our study are likely skewed toward EUR ancestry, as was evidenced by the trivial differences between the predictive performance of PGSs based on EUR-like ancestry-specific and trans-ancestry GWASs. Also, the genetic associations across ancestral groups may be influenced by reduced coverage of common genetic variants in non-EUR-like groups^67^. The non-EUR-like ancestry groups were considerably smaller and had less power for ancestry-specific GWAS, post-GWAS analyses, or for computing cross-ancestry PGSs by meta-analyzing ancestry-specific results with PRS-CSx^68^.

Nevertheless, the increased number of genomic risk loci discovered upon including the non-EUR-like participants demonstrates the advantages of our ancestrally inclusive pipeline. Moreover, the non-EUR-like ancestry-specific results from this study may contribute to large-scale ancestry-specific and cross-ancestry GWAS meta-analyses of MDD. To achieve parity in the population-specific understanding of the genetics of MDD, continued long-term efforts to recruit currently underrepresented populations remain imperative. Likewise, it is essential to build globally representative reference datasets to facilitate post-GWAS analyses^69^.

Overall, trans-ancestry GWAS of depression outcomes maximized the genomic insights from extant data and uncovered novel genomic risk loci for MDD. Our findings extend the prior genome-level findings^17^ of little overlap between MDD and broad depression to genomic locus and gene expression levels, re-emphasizing the need for investing in larger, globally representative datasets with strictly defined, or preferably clinically evaluated, MDD. Such studies will be vital for bridging the gaps in our understanding of MDD and developing novel disease-specific therapies generalizable across populations. In the immediate term, our analytical pipeline may facilitate further trans-ancestry analyses of existing data to help improve global representation, statistical power, and generalizability in genomic studies.

## Data Availability

The UK Biobank data used in this research is available to “bona fide researchers for health-related research in the public interest” through an application process accessible through the UK Biobank website, https://www.ukbiobank.ac.uk/.

The summary statistics from the genome-wide association analyses will be made available through OSF.

## Acknowledgments

This research has been conducted using the UK Biobank Resource application number 30782 (PI: REP). We gratefully acknowledge Dr. Kenneth S. Kendler for his valuable guidance and feedback on the MDD phenotypes derived *in silico* and analyzed in this study. The authors wish to acknowledge their respective consortia and networks including the PGC Cross-Population Analyses Working Group (REP, MS, CC, TBB, AEG, BTW; https://pgc.unc.edu/for-researchers/working-groups/cross-population-analyses-working-group/), and the PsycheMERGE Network Diversity Initiative (REP, MS, CC, PBB, TBB; https://psychemerge.com/diversity-initiative-workgroup/). This work was supported by the National Institutes of Health grants R01MH125938 (REP, MS, CC, AEG, TBB, BTW) and K01AA031748 (AEG) and the Brain Behavior Research Foundation NARSAD grant 28632 P&S Fund (REP). Computing costs in the All of Us Researcher Workbench were supported by R01MH125938 (PI: REP). The content is solely the responsibility of the authors and does not necessarily represent the official views of the funding agencies and the National Institutes of Health.

